# Effect of Adjunctive Inhalation on the Association Between Plasma AUC/MIC of Polymyxin B and Clinical Efficacy in MDR Gram-Negative Infections

**DOI:** 10.64898/2026.04.29.26352086

**Authors:** Shengnan Zhang, Yaqian Li, Hongyi Tan, Yejun Li, Yuanfang Qin, Tingting Wu, Jingjing Liu, Qi Pei

**Author notes:** Corresponding author: Qi Pei,; The Third Xiangya Hospital, Central South University, Yuelu District, Changsha, Hunan 410013, People’s Republic of China., Jingjing Liu,; The Third Xiangya Hospital, Central South University, Yuelu District, Changsha, Hunan 410013, People’s Republic of China. Shengnan Zhang and Yaqian Li contributed equally to this work.

## Abstract

**Objectives:** To develop a population pharmacokinetic (PPK) model of polymyxin B (PMB) for intravenous (IV) and combined intravenous plus inhaled (IV+IH) administration in critically ill patients, and evaluate the association between the 24-h steady-state area under concentration–time curve to minimum inhibitory concentration ratio (AUC_ss,24h_/MIC) and clinical outcomes.

**Methods:** This prospective cohort was conducted in the ICU of the Third Xiangya Hospital, Central South University (ethics R19048; ChiCTR1900028602). Adults with multidrug-resistant Gram-negative bacterial infections receiving PMB ≥48 h were enrolled and assigned to IV or IV+IH groups. Serial plasma samples were analyzed by validated LC–MS/MS. The PPK model was developed with NONMEM^®^. Clinical efficacy at end of treatment was blindly assessed.

**Results:** Forty-three patients were enrolled (IV, n=22; IV+IH, n=21), with an overall clinical success rate of 66.7%. A two-compartment PPK model best described the data, with typical values of clearance (2.6 L/h), central volume (13.6 L), and peripheral volume (17.6 L). Clearance was influenced by creatinine clearance and total bile acids. In the overall cohort, neither AUC_ss,24h_ nor AUC_ss,24h_/MIC differed significantly between clinical success and failure (*p*=0.591 and 0.143). In the IV group, AUC_ss,24h_/MIC was significantly higher in responders (*p*=0.005) with an ROC-derived efficacy threshold of 94.37; AUC_ss,24h_ showed a non-significant trend (*p*=0.076). No exposure– response relationship was observed in the IV+IH group (*p*=0.398 and 0.495).

**Conclusions:** Plasma AUC_ss,24h_/MIC appears to be associated with clinical efficacy during IV monotherapy but not in IV+IH regimens, likely due to high pulmonary exposure. Plasma-based PK/PD targets should be applied cautiously when inhalation is added.

## 1. Introduction

The global crisis of multidrug-resistant gram-negative bacteria (MDR-GNB) infections continues to intensify while current therapeutic strategies face formidable challenges. In the 2024 WHO priority pathogen list, carbapenem-resistant *Acinetobacter baumannii*, Enterobacterales, and third-generation cephalosporin-resistant Enterobacterales are designated “critical priority,” and carbapenem-resistant *Pseudomonas aeruginosa* is designated “high priority” ^1^. Epidemiological studies indicate that hospital-acquired pneumonia and ventilator-associated pneumonia caused by these pathogens carry overall mortalities as high as 40–53% ^2–4^. Against this backdrop, rational use of polymyxin B—one of the last-line antibacterial agents—has become essential. Yet its clinical application faces a dual dilemma: systemic high-dose therapy is limited by nephrotoxicity and neurotoxicity ^5–7^, while intravenous administration yields suboptimal penetration into lung tissues. Studies have reported an epithelial lining fluid (ELF)–to-plasma concentration ratio of only 15.7% ^8^, potentially leading to unsatisfactory outcomes in pulmonary infections.

To address this limitation, combined intravenous administration (IV) and inhaled PMB therapy has garnered attention. By delivering drug directly to the lungs, inhalation can markedly increase local concentrations ^9–11^. The International Consensus Guidelines ^12^ recommend adjunctive inhaled polymyxin for suspected or confirmed extensively drug resistant-pathogen hospital acquired pneumonia/ventilator associated pneumonia when IV PMB is required, and suggest a steady-state 24-hour area under the concentration–time curve (AUC_ss,24h_) of 50–100 mg·h/L as a PK/PD target to guide dosing. Notably, this target was extrapolated from thigh-infection PK/PD studies in mice receiving IV polymyxin ^13,14^, and the relationship between AUC/MIC and efficacy has not been verified for combined IV plus inhalation (IV+IH) regimens. Given the increasing adoption of combined therapy, re-evaluating the suitability of this target is warranted.

Most PPK studies of PMB have focused on IV administration, whereas the pharmacokinetics of combined IV plus inhalation remain insufficiently characterized ^15–17^. This study aimed to construct a PMB PPK model covering both routes to describe systemic exposure following IV and IV+IH dosing, and thereby to assess whether plasma AUC/MIC correlates with clinical efficacy under each route—particularly whether, under IV+IH where high local pulmonary exposure may dominate, the predictive power of plasma AUC/MIC weakens or disappears.

## 2. Methods

### 2.1 Study design and ethics

This prospective observational cohort study was conducted in the Department of Critical Care Medicine, the Third Xiangya Hospital of Central South University from January 2019 to December 2020. The protocol was approved by the Ethics Committees of the Third Xiangya Hospital of Central South University (R19048) and registered at the Chinese Clinical Trial Registry (ChiCTR1900028602). Written informed consent was obtained from all participants or their legal representatives. The study complied with the Declaration of Helsinki and Good Clinical Practice.

### 2.2 Patient enrollment and exclusion

Inclusion criteria were: (i) age ≥18 years; (ii) MDR-GNB infection confirmed according to CLSI 2019 susceptibility standards; (iii) PMB treatment ≥48 h; treatment regimens comprised IV monotherapy or combined IV plus inhalation. Exclusion criteria were: (i) concomitant use of other polymyxins; (ii) severe drug allergy history; (iii) pregnancy or lactation. Demographic characteristics and routine laboratory data were retrieved from the electronic medical record.

### 2.3 Dosing regimens and sample collection

Intravenous administration: loading dose (if applicable) 100–150 mg; maintenance 50–100 mg q12h, adjusted by clinicians based on body weight, renal function, and infection severity; infusion duration 0.5–1 h. Combined regimen: PMB 25 mg by inhalation plus 75 mg IV infusion q12h. Inhalation employed a vibrating-mesh nebulizer (GENTEC, GUN-300VT) for ∼15 min. At steady state (defined as ≥4 consecutive doses of the same regimen), venous blood was collected at pre-dose and 1, 3, 5, 9, and 12 h post-dose. Samples were centrifuged and stored at −80 °C.

### 2.4 PMB concentration assay

An analytically validated HPLC–MS/MS method quantified PMB. Chromatography used an Agilent Eclipse XDB-C18 column (2.1×150 mm, 3.5 μm). Mobile phases were 0.3% formic acid in water (A) and 0.3% formic acid in acetonitrile (B), with gradient elution at 0.4 mL/min. Mass spectrometry employed positive-ion ESI; quantifier transitions were m/z 602.7→241.3 for PMB1 and 595.5→227.1 for PMB2. Polymyxin E2 served as internal standard (m/z 578.1→101.1). Linearity ranges were 0.05–10 mg/L for PMB1 and 0.011–1.097 mg/L for PMB2. Total PMB concentration was the sum of PMB1 and PMB2. Intra-/inter-day precision (CV ≤12.2%) and accuracy (88.1–114.4%) met acceptance criteria.

### 2.5 Population pharmacokinetic modeling and validation

NONMEM^®^ (version 7.3; ICON Development Solutions, Ellicott City, MD, USA) was used to analyze concentration–time data, with FOCEI estimation. One- and two-compartment structural models were compared. Model selection considered decreases in objective function value (OFV), visual inspection of goodness-of-fit (GOF) plots, and pharmacological plausibility. Interindividual variability (IIV) was modeled exponentially. Residual variability models (additive, proportional, combined) were evaluated. Stepwise covariate modeling assessed effects of demographics, renal function, inflammatory markers, and hepatic indices on PK parameters (forward inclusion *p*<0.05; backward elimination *p*<0.001). Internal validation included: (1) GOF plots; (2) nonparametric bootstrap (n=1000); and (3) prediction-corrected visual predictive checks (pcVPC; n=1000 simulations). Post-processing and visualization were performed in R (version 4.3.1; http://www.r-project.org).

### 2.6 Exposure metrics and MIC determination

Using the final PPK model, Bayesian posterior estimates provided individual AUC_ss,24h_. Pathogens were isolated from relevant infection sites; MICs were determined with the VITEK-2 Compact system (bioMérieux, France). AUC_ss,24h_/MIC, calculated using the MIC from the most recent pre-treatment isolate, served as the key PK/PD index for efficacy prediction. If multiple potential pathogens were isolated, the isolate best matching the clinical infection focus was prioritized.

### 2.7 Efficacy assessment and statistical analysis

The primary endpoint was clinical efficacy at end of therapy, blindly assessed by two infectious-disease experts not involved in patient care based on symptoms, signs, and laboratory results: success (cure: complete resolution of clinical signs/symptoms with normalization of labs; or clinical improvement: marked improvement from baseline) versus failure (persistence or progression: no improvement or worsening; or all-cause death). Secondary endpoints included microbiological eradication and ICU length of stay. To minimize assessment bias, all information related to treatment modality (route, dose, concomitant therapy) was concealed. Discrepancies were resolved by a third adjudicator or consensus. Statistical analyses used R 4.3.1. Wilcoxon rank-sum tests compared AUC_ss,24h_, AUC_ss,24h_/MIC, and trough concentrations between success and failure. ROC curves explored relationships between PK indices and efficacy. For baseline comparisons, continuous variables were analyzed by Student’s t-test or Mann–Whitney U test as appropriate; categorical variables were analyzed by chi-square or Fisher’s exact test. Two-sided *α*=0.05; *p*<0.05 indicated statistical significance.

## 3. Results

### 3.1 Patient characteristics

A total of 43 ICU patients were enrolled, including 33 males; mean age 59.63 ± 17.50 years and mean body weight 65.93 ± 14.76 kg. 21 received IV+IH and 22 received IV alone. Daily PMB doses ranged from 100–300 mg/day; 88.4% received 100 mg q12h. Treatment duration was 3–24 days. Hospital length of stay ranged from 8–142 days; ICU stay from 8–71 days. Detailed demographics and treatment data are shown in Table 1.

**Table 1.**
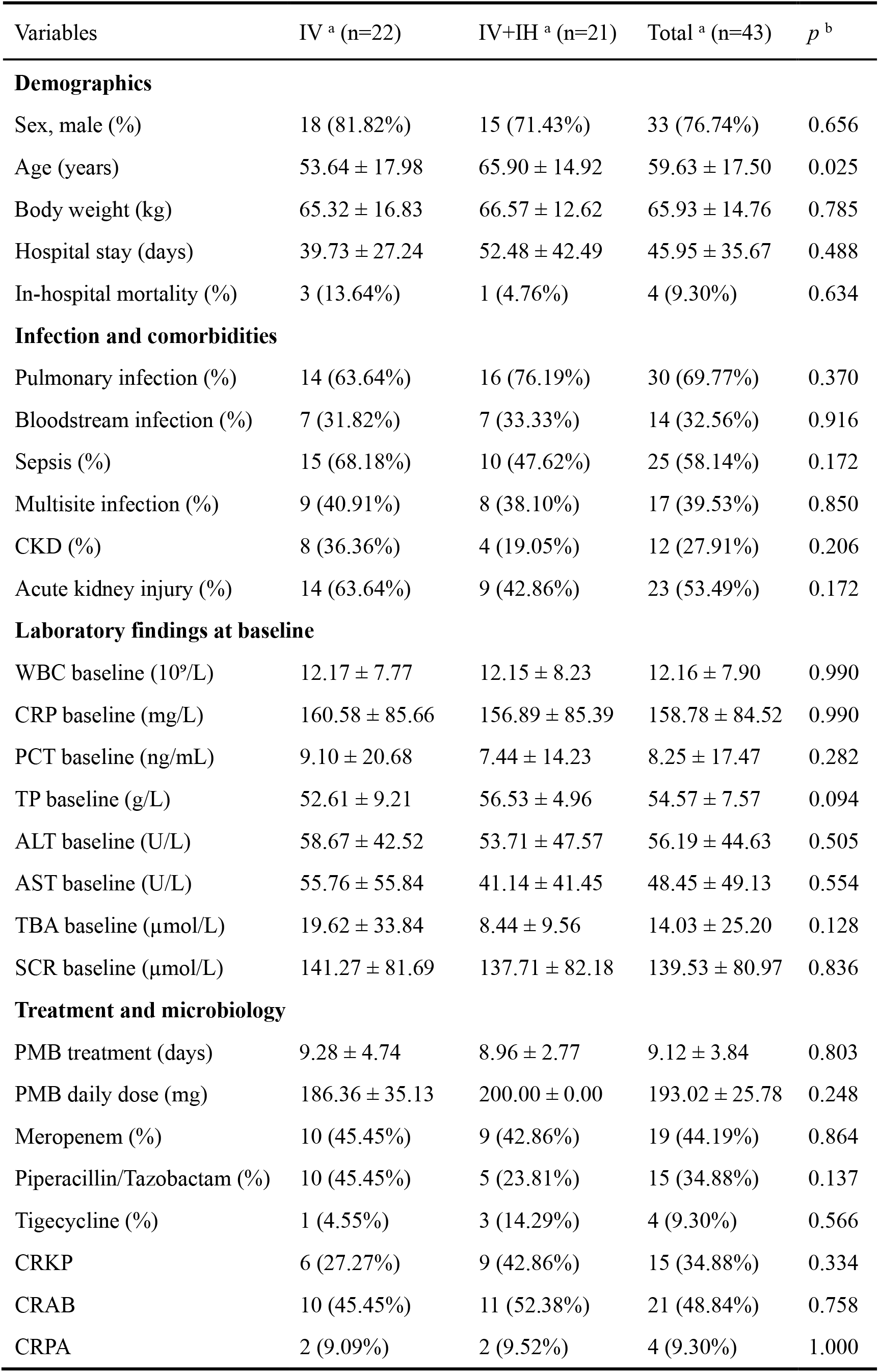
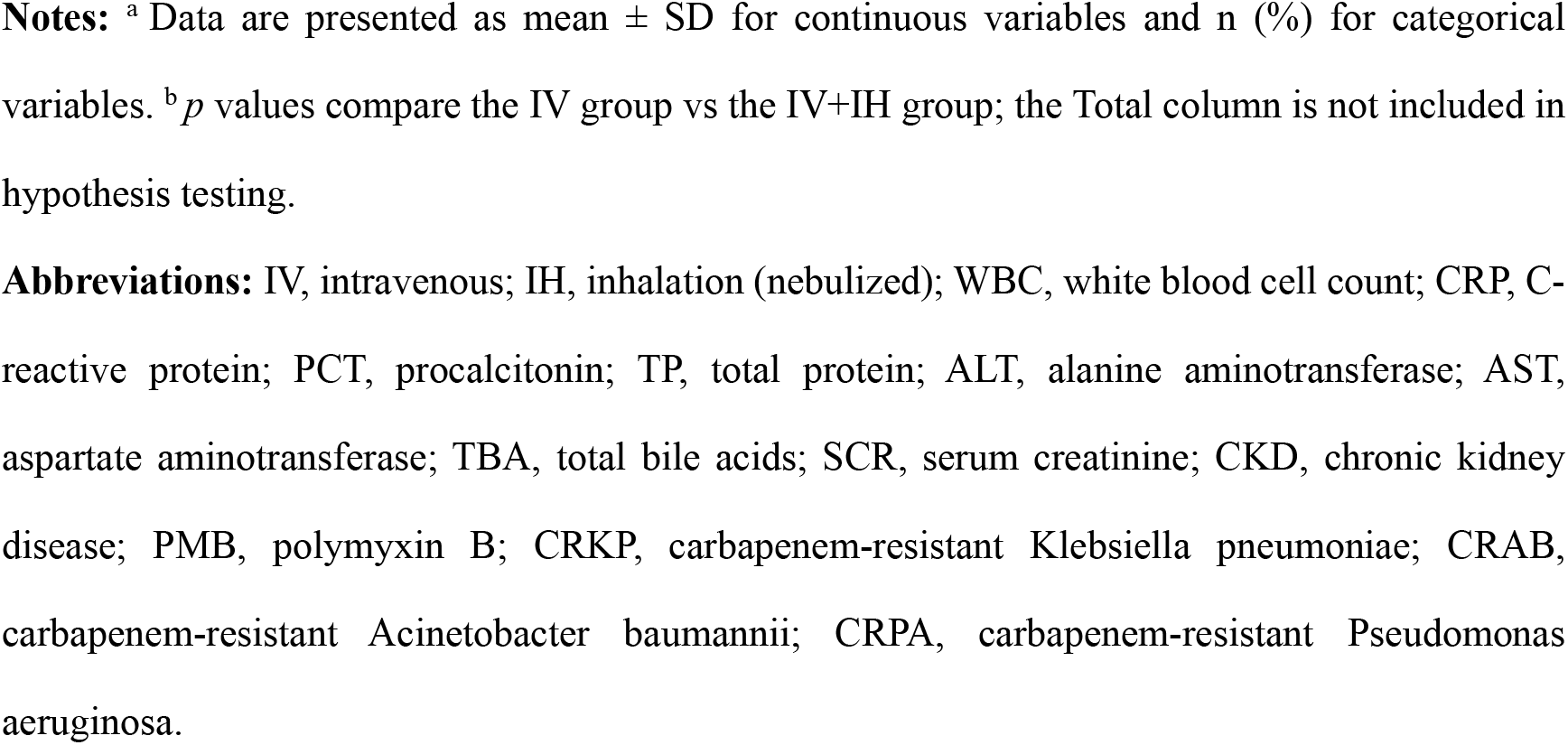
Baseline demographics, comorbidities, laboratory findings, and treatment characteristics stratified by dosing regimen.

| Variables | IV <sup>a</sup> (n=22) | IV+IH <sup>a</sup> (n=21) | Total <sup>a</sup> (n=43) | *p* <sup>b</sup> |
| --- | --- | --- | --- | --- |
| **Demographics** |  |  |  |  |
| Sex, male (%) | 18 (81.82%) | 15 (71.43%) | 33 (76.74%) | 0.656 |
| Age (years) | 53.64 ± 17.98 | 65.90 ± 14.92 | 59.63 ± 17.50 | 0.025 |
| Body weight (kg) | 65.32 ± 16.83 | 66.57 ± 12.62 | 65.93 ± 14.76 | 0.785 |
| Hospital stay (days) | 39.73 ± 27.24 | 52.48 ± 42.49 | 45.95 ± 35.67 | 0.488 |
| In-hospital mortality (%) | 3 (13.64%) | 1 (4.76%) | 4 (9.30%) | 0.634 |
| **Infection and comorbidities** |  |  |  |  |
| Pulmonary infection (%) | 14 (63.64%) | 16 (76.19%) | 30 (69.77%) | 0.370 |
| Bloodstream infection (%) | 7 (31.82%) | 7 (33.33%) | 14 (32.56%) | 0.916 |
| Sepsis (%) | 15 (68.18%) | 10 (47.62%) | 25 (58.14%) | 0.172 |
| Multisite infection (%) | 9 (40.91%) | 8 (38.10%) | 17 (39.53%) | 0.850 |
| CKD (%) | 8 (36.36%) | 4 (19.05%) | 12 (27.91%) | 0.206 |
| Acute kidney injury (%) | 14 (63.64%) | 9 (42.86%) | 23 (53.49%) | 0.172 |
| **Laboratory findings at baseline** |  |  |  |  |
| WBC baseline (10<sup>9</sup>/L) | 12.17 ± 7.77 | 12.15 ± 8.23 | 12.16 ± 7.90 | 0.990 |
| CRP baseline (mg/L) | 160.58 ± 85.66 | 156.89 ± 85.39 | 158.78 ± 84.52 | 0.990 |
| PCT baseline (ng/mL) | 9.10 ± 20.68 | 7.44 ± 14.23 | 8.25 ± 17.47 | 0.282 |
| TP baseline (g/L) | 52.61 ± 9.21 | 56.53 ± 4.96 | 54.57 ± 7.57 | 0.094 |
| ALT baseline (U/L) | 58.67 ± 42.52 | 53.71 ± 47.57 | 56.19 ± 44.63 | 0.505 |
| AST baseline (U/L) | 55.76 ± 55.84 | 41.14 ± 41.45 | 48.45 ± 49.13 | 0.554 |
| TBA baseline (μmol/L) | 19.62 ± 33.84 | 8.44 ± 9.56 | 14.03 ± 25.20 | 0.128 |
| SCR baseline (μmol/L) | 141.27 ± 81.69 | 137.71 ± 82.18 | 139.53 ± 80.97 | 0.836 |
| **Treatment and microbiology** |  |  |  |  |
| PMB treatment (days) | 9.28 ± 4.74 | 8.96 ± 2.77 | 9.12 ± 3.84 | 0.803 |
| PMB daily dose (mg) | 186.36 ± 35.13 | 200.00 ± 0.00 | 193.02 ± 25.78 | 0.248 |
| Meropenem (%) | 10 (45.45%) | 9 (42.86%) | 19 (44.19%) | 0.864 |
| Piperacillin/Tazobactam (%) | 10 (45.45%) | 5 (23.81%) | 15 (34.88%) | 0.137 |
| Tigecycline (%) | 1 (4.55%) | 3 (14.29%) | 4 (9.30%) | 0.566 |
| CRKP | 6 (27.27%) | 9 (42.86%) | 15 (34.88%) | 0.334 |
| CRAB | 10 (45.45%) | 11 (52.38%) | 21 (48.84%) | 0.758 |
| CRPA | 2 (9.09%) | 2 (9.52%) | 4 (9.30%) | 1.000 |

**Notes:** <sup>a</sup> Data are presented as mean ± SD for continuous variables and n (%) for categorical variables. <sup>b</sup> *p* values compare the IV group vs the IV+IH group; the Total column is not included in hypothesis testing.
**Abbreviations:** IV, intravenous; IH, inhalation (nebulized); WBC, white blood cell count; CRP, C-reactive protein; PCT, procalcitonin; TP, total protein; ALT, alanine aminotransferase; AST, aspartate aminotransferase; TBA, total bile acids; SCR, serum creatinine; CKD, chronic kidney disease; PMB, polymyxin B; CRKP, carbapenem-resistant *Klebsiella pneumoniae*; CRAB, carbapenem-resistant *Acinetobacter baumannii*; CRPA, carbapenem-resistant *Pseudomonas aeruginosa*.

### 3.2 Final model

A total of 208 PMB plasma concentrations from 43 critically ill patients were included in the population pharmacokinetic model development. Patient demographics and baseline clinical characteristics are presented in Table 1. Compared with the one-compartment structural model, the two-compartment model provided a lower OFV and reduced systematic bias in diagnostic plots. The final model was a two-compartment pharmacokinetic model incorporating first-order absorption for the inhalation route (Figure 1). A combined proportional and additive residual error model best described the residual variability. Creatinine clearance (CRCL) and total bile acids (TBA) were identified as significant covariates on clearance (CL). Population parameter estimates (Table 2) were: typical CL, 2.6 L/h; central volume of distribution (V_C_), 13.6 L; peripheral volume of distribution (V_P_), 17.6 L; and intercompartmental clearance (Q), 22.1 L/h. IIV was estimated at 35.9% for CL and 67.2% for V_C_. GOF plots (Figure 2) indicated satisfactory overall fit, with data points distributed around the line of identity (y=x). Conditional weighted residuals versus time and predictions were symmetrically scattered around zero, without discernible trends. The VPC plot (Figure 3) confirmed the predictive performance of the model, with the observed 5th, 50th, and 95th percentiles falling within the 95% prediction intervals generated from 1000 simulations. Bootstrap analyses demonstrated robustness of the final model, with >80% successful minimizations and parameter estimates fell within the bootstrap-derived 95% confidence intervals.

**Table 2.** Population PK parameter estimates of final model and bootstrap validation.

| Parameter | Final model |  | Bootstrap |  |
| --- | --- | --- | --- | --- |
|  | Estimate | 95% CI | Estimate | 95% CI |
| CL (L/h) | 2.61 | 2.18 - 3.04 | 2.61 | 2.22 - 3.00 |
| K<sub>a</sub> (h<sup>-1</sup>) | 0.018 | 0.001 - 0.050 | 0.026 | 0.003 - 1.064 |
| V<sub>c</sub> (L) | 13.59 | 6.03 - 21.17 | 14.86 | 7.87 - 21.68 |
| V<sub>p</sub> (L) | 17.58 | 10.92 - 24.28 | 17.89 | 12.37 - 32.34 |
| Q (L/h) | 21.17 | 6.72 - 35.68 | 17.06 | 5.87 - 28.60 |
| CL_CRCL | 0.239 | 0.097 - 0.381 | 0.261 | 0.055 - 0.424 |
| CL_TBA | 0.112 | 0.062 - 0.162 | 0.124 | 0.047 - 0.199 |
| Between-subject variability ($\omega^2$) | | | | |
| CL | 0.129 |  | 0.129 |  |
| V<sub>c</sub> | 0.452 |  | 0.342 |  |
| Residual error ($\sigma^2$) | | | | |
| Proportional | 0.026 |  | 0.016 |  |
| Additive | 0.006 |  | 0.009 |  |
| Bootstrap success rate | 86% |  |  |  |

**Abbreviations:** CI, confidence interval; CL, clearance; K<sub>A</sub>, first-order absorption rate constant (for inhalation); V<sub>C</sub>, central volume of distribution; V<sub>P</sub>, peripheral volume of distribution; Q, intercompartmental clearance; CL\_CRCL and CL\_TBA, covariate exponents on CL for creatinine clearance and total bile acids, respectively; $\omega^2$ , between-subject variance; $\sigma^2$ , residual variance.

**Figure 1.**
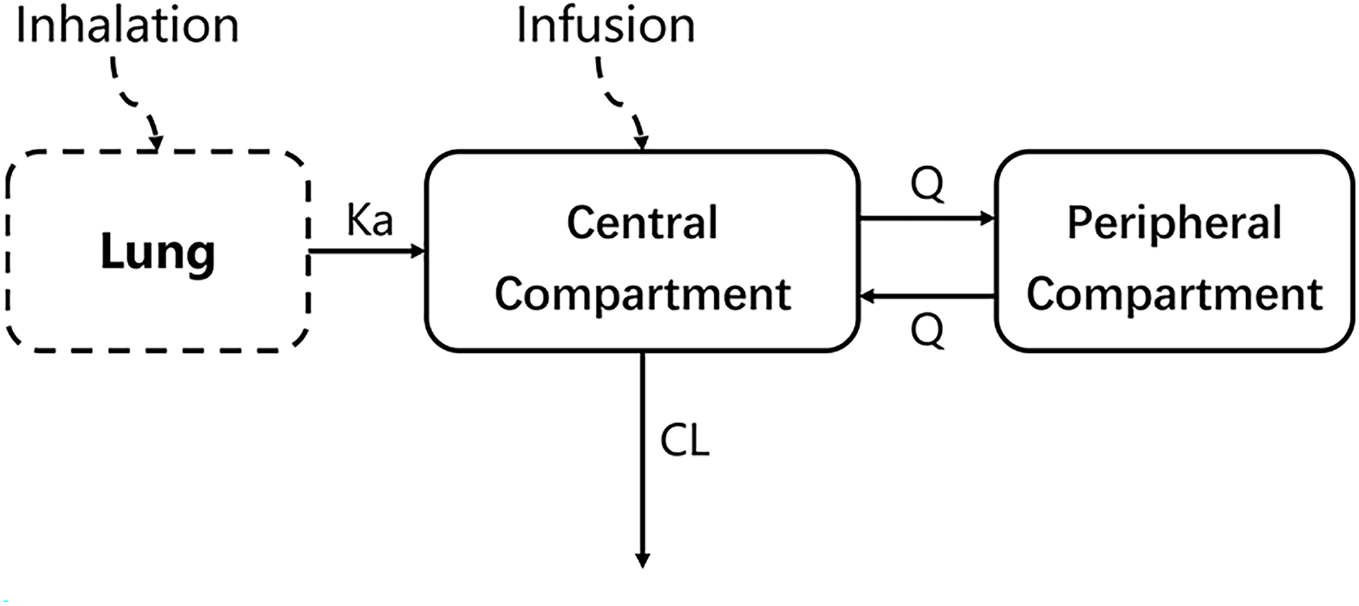
Final population PK structural model

**Figure 2.**
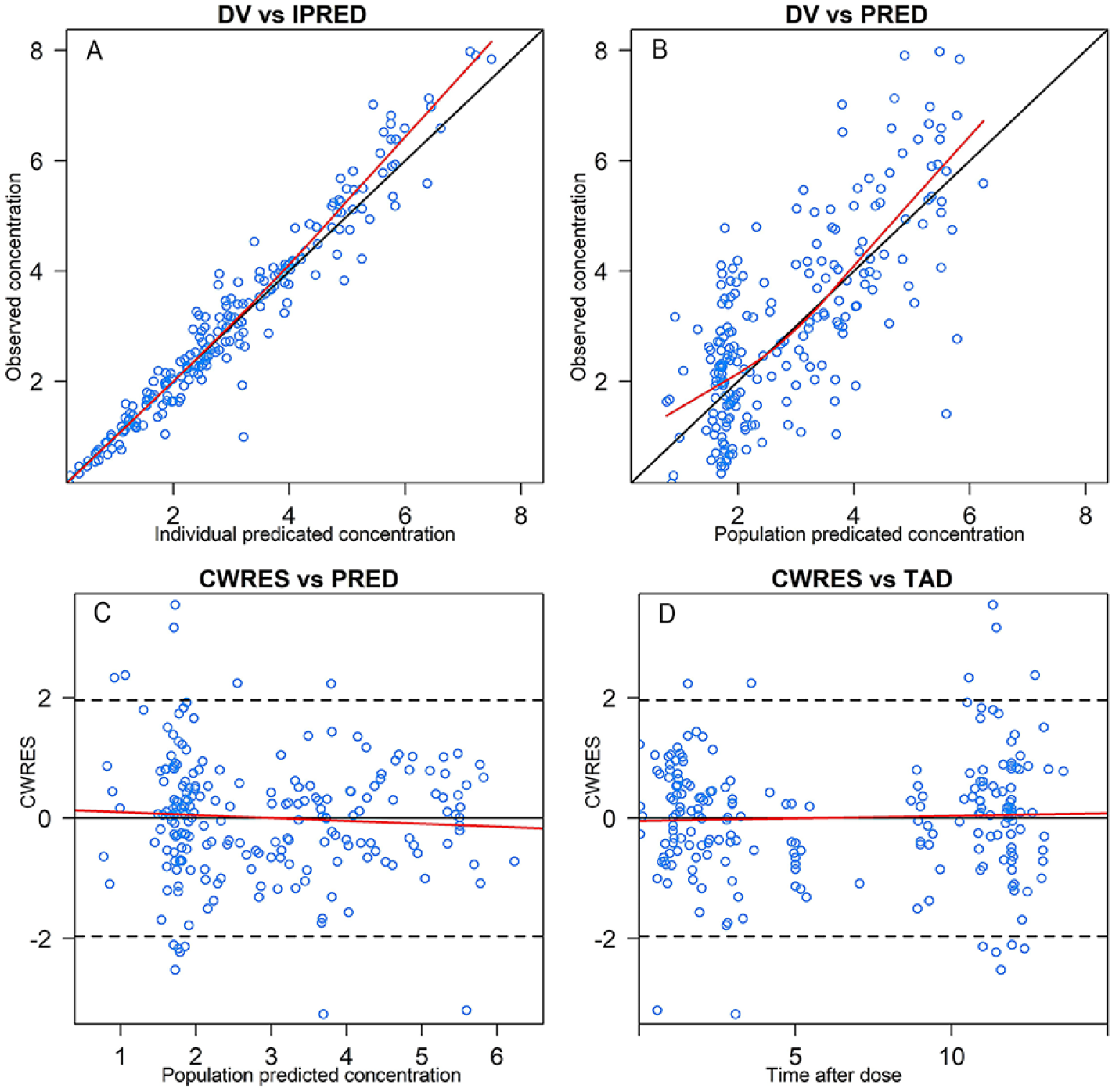
Goodness-of-fit diagnostic plots for the final population PK model. **Notes:** Panels A–D correspond to DV vs IPRED, DV vs PRED, CWRES vs PRED, and CWRES vs time.

**Figure 3.**
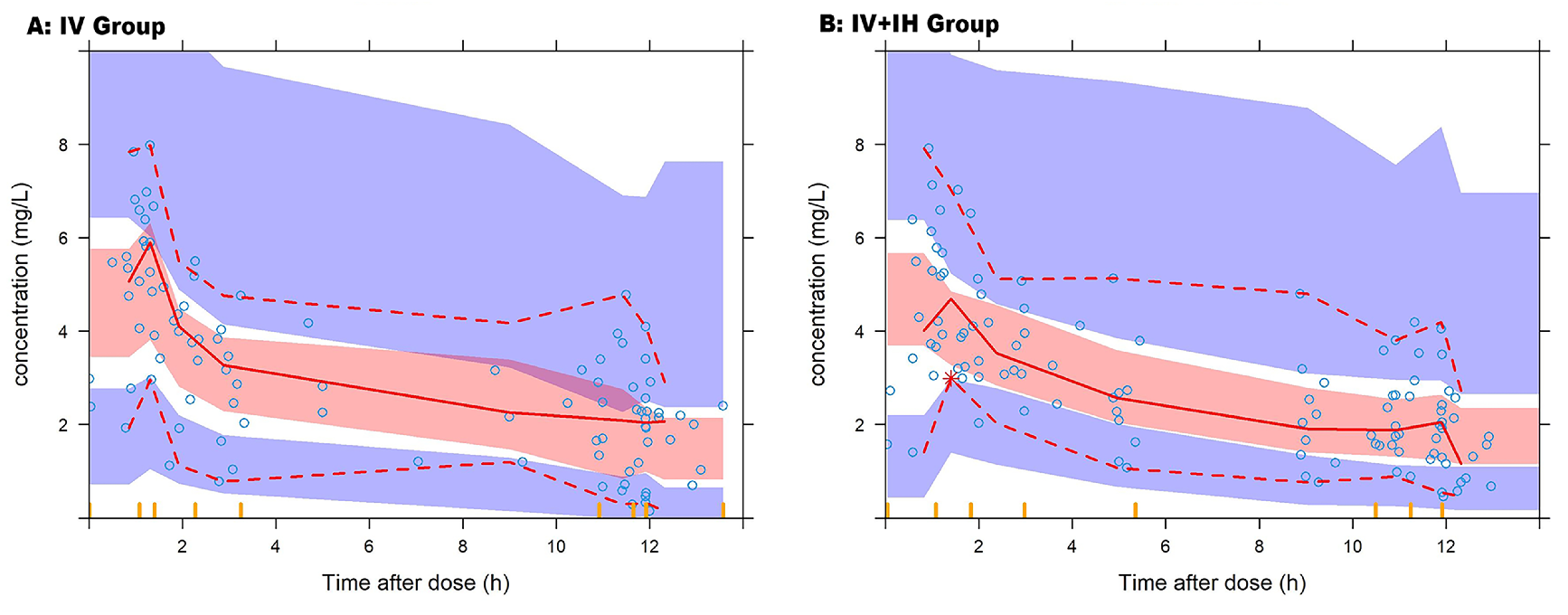
Prediction-corrected visual predictive check. **Notes:** (A) Intravenous (IV) only; (B) Combined intravenous plus inhalation (IV+IH). Points denote observed plasma concentrations (mg/L) versus time after dose (h); solid lines show the observed 5th, 50th, and 95th percentiles; shaded ribbons indicate the 95% prediction intervals of the corresponding simulated percentiles.

### 3.3 Efficacy –response analysis

Efficacy was assessed in 42 critically ill patients (one patient in IV group was excluded due to missing pathogen culture data). Overall, 28 patients (66.7%) achieved clinical success, whereas 14 patients (33.3%) experienced treatment failure. In the entire cohort (Figure 4), neither AUC_ss,24h_ nor AUC_ss,24h_/MIC demonstrated a statistically significant difference between patients with clinical success and failure (*p*=0.591 and *p*=0.143, respectively). Subgroup analysis was subsequently performed by treatment regimen, dividing patients into IV group and IV+IH group. The clinical success rate was 66.7% in both groups. In the IV subgroup, clinical efficacy was associated with plasma pharmacokinetic exposure. Both AUC_ss,24h_ and AUC_ss,24h_/MIC were higher among responders, with AUC_ss,24h_/MIC reaching statistical significance (*p*=0.005) and AUC_ss,24h_ showed a non-significant trend (*p*=0.076) (Figure 4). ROC analysis identified an AUC_ss,24h_/MIC threshold of 94.37 for predicting clinical success (AUC 0.740; sensitivity 0.692; specificity 0.875; J=0.567). By contrast, in the IV+IH subgroup, no significant association was observed between clinical efficacy and either AUC_ss,24h_ or AUC_ss,24h_/MIC (*p*=0.398 and *p*=0.495, respectively).

**Figure 4.**
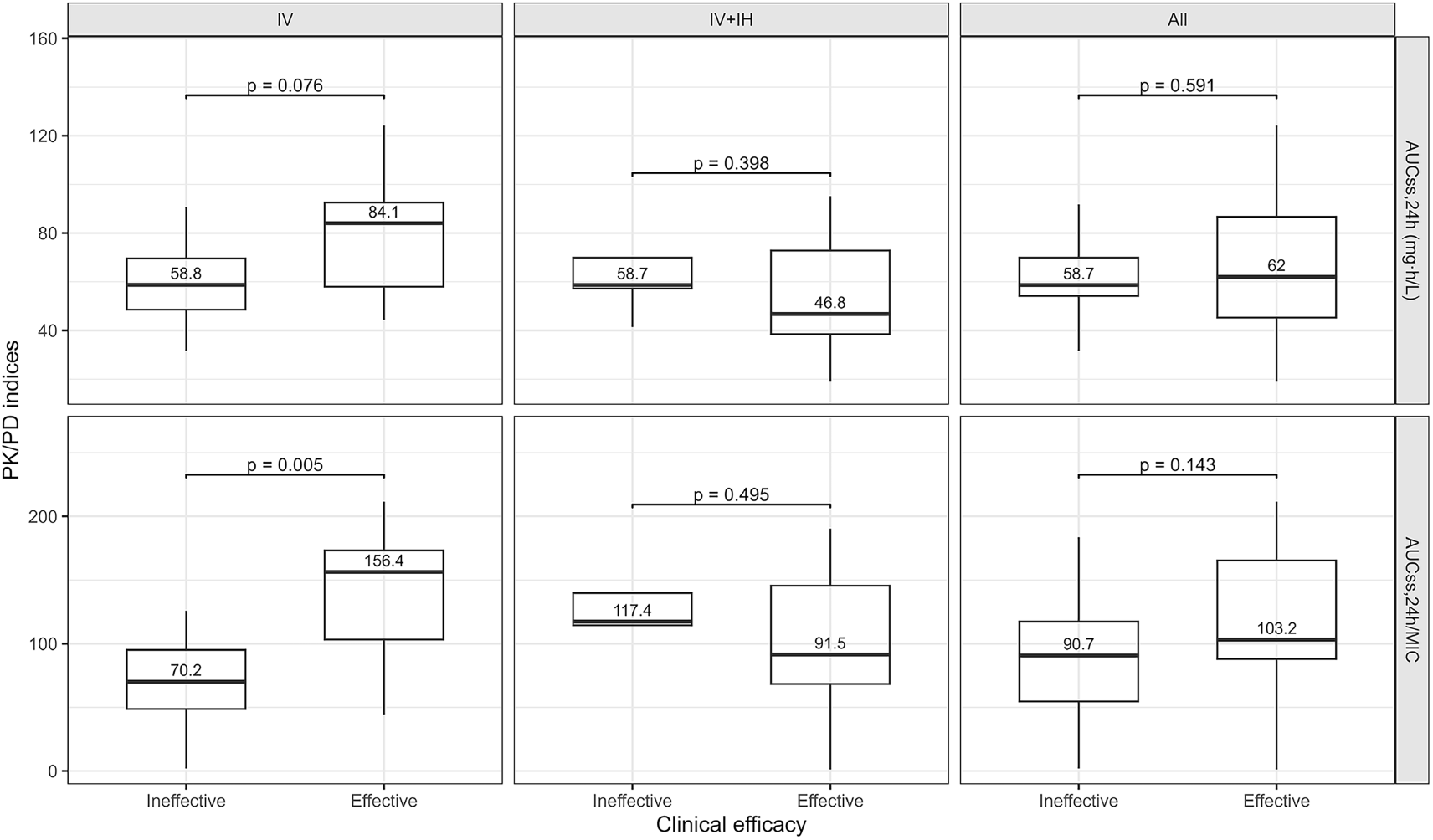
Exposure–response analyses stratified by dosing route. **Abbreviations:** IV, Intravenous only; IV+IH, Combined intravenous plus inhalation therapy

## 4. Discussion

As threats from Gram-negative pathogens—particularly MDR organisms such as *Klebsiella pneumoniae* and *A. baumannii*—have escalated over recent decades, the clinical role of polymyxin B as a “last-line” antibiotic has been re-established ^18^. Our PPK model yielded a typical CL of 2.6 L/h (IIV 35.9%), close to the median 2.43 L/h (IIV 29.8%) reported in a systematic review ^19^. The median central volume (13.6 L) also approximated the reported 13.4 L ^19^, supporting model reliability. The substantial IIV observed for CL and V_C_ aligns with pathophysiological features common in critical illness—capillary leak, fluid shifts, hypoalbuminemia, and fluctuating organ perfusion. We identified CRCL and TBA as significant covariates on CL. Although PMB is primarily cleared non-renally with low urinary recovery (<5%), and guidelines emphasize weak theoretical dependence on renal function, numerous models have nonetheless found CRCL to be a significant covariate on CL ^20–24^. In our data, TBA correlated positively with CL, albeit with a modest effect (power exponent 0.112). Elevated TBA may reflect altered liver function or bile acid homeostasis, potentially influencing polymyxin B clearance indirectly via effects on transporters or plasma protein binding ^25,26^. Given the modest effect and limited mechanistic evidence, its clinical significance requires confirmation in future studies.

From a pharmacodynamic perspective, the free drug AUC/MIC (*f*AUC/MIC) is considered the principal PK/PD index of polymyxin antimicrobial activity ^13,27–31^. In a murine thigh model, Li et al. reported that a median plasma *f*AUC/MIC of 10.0 achieved a 1-log reduction in across all three organisms^14^. Clinically, total-drug AUC/MIC—more readily monitored—has been associated with therapeutic outcomes ^32–34^. Based on animal data and clinical observations, and assuming a plasma protein binding of 58% (unbound 42%), an AUC_ss,24h_ target range of 50–100 mg·h/L is commonly adopted for Gram-negative infections with MIC of 2 mg/L ^14,30^. However, the applicability of this systemic target to pulmonary infections—especially with adjunctive inhalation—has been challenged. In a murine pneumonia model, Li et al. demonstrated that an *f*AUC/MIC of at least 43.3 was required to achieve a 1-log10 bacterial reduction in lung tissue, a value substantially higher than that observed in the thigh-infection model ^30^. In our study, AUC_ss,24h_/MIC was significantly associated with efficacy in the IV-only subgroup, but this association was lost in both the IV+IH subgroup and the overall cohort. Similarly, Tang et al. ^34^ in a study of 105 patients with carbapenem-resistant *A. baumannii*, identified AUC_ss,24h_/MIC as an important determinant of efficacy, with an ROC-derived optimal cutoff of approximately 66.9.

One plausible explanation is that, inhalation delivers PMB particles directly to the infected lung, forming a high-concentration drug layer over alveolar and bronchial mucosa, effectively penetrating biofilms and achieving local exposures far exceeding those attainable with IV dosing alone ^9–11,35^. Therapeutic drug monitoring has documented ELF PMB peaks up to 116.7 mg/L in patients receiving combined therapy ^10^, vastly higher than contemporaneous plasma concentrations typically in the low single-digit mg/L range, and sufficient to exceed MICs for most common pathogens for prolonged periods. In contrast, the highly polar and large molecular structure of PMB restricts its tissue distribution, resulting in suboptimal pulmonary penetration with IV monotherapy ^8,18^.

Administration via inhalation can circumvent this limitation by delivering high local concentrations directly to the lung. Furthermore, as inhaled doses are relatively small and systemic absorption is limited, plasma exposure increases only modestly ^11,35^, thereby potentially enhancing pulmonary efficacy without a corresponding rise in systemic toxicity.

This single-center, prospective observational study has methodological and interpretive limitations. First, the total sample size and subgroup sizes were small; statistical power was limited for IV versus IV+IH analyses, ROC cutoffs may be unstable, and type-II error is possible. Second, we did not collect ELF or other pulmonary surrogate samples and therefore could not quantify the contribution of inhalation to local lung exposure; mechanistic inferences relied on prior literature and physiological plausibility. Finally, MICs were measured using an automated commercial susceptibility system. Compared with the broth microdilution gold standard, automated systems may underestimate polymyxin MICs, potentially under-appreciating resistance. As emphasized by Yang et al.^32^, accuracy of MIC determination warrants heightened attention and caution when optimizing doses using PK/PD principles.

## 5. Conclusions

Based on a PPK model encompassing IV and IV+IH dosing, our findings suggest that the suitability of plasma AUC/MIC as a surrogate efficacy marker in severe MDR-GNB infections depends on the route of administration. Among patients receiving IV monotherapy, higher AUC_ss,24h_/MIC was significantly associated with clinical success. In contrast, among those receiving combined IV plus inhalation, the association between plasma AUC/MIC and efficacy was no longer significant. These observations challenge current dosing-optimization strategies centered on plasma AUC/MIC and provide a rationale for mechanistic studies and outcome evaluations specific to inhalation therapy.

## Data Availability

All data produced in the present study are available upon reasonable request to the authors

## Author contributions

QP and JJL contributed to the conception and design of the study. QP performed data analysis, modeling, secured funding, and supervised the project. JJL contributed to study design, data collection, data analysis, and supervision. YQL and SNZ were responsible for data collection, modeling, analysis, visualization, and drafting of the manuscript. HYT contributed to study design, sample determination, and data analysis. YJL performed sample determination and data analysis. TTW and YFQ contributed to data collection and analysis. All authors reviewed and approved the final version of the manuscript.

## Ethics statement

This study was approved by the Ethics Committee of the Third Xiangya Hospital of Central South University (approval number: R19048) and registered in the Chinese Clinical Trial Registry (ChiCTR1900028602).

## Data availability

Data will be made available on reasonable request.

